# Evaluation of COVID-19 vaccine breakthrough infections among immunocompromised patients fully vaccinated with BNT162b2

**DOI:** 10.1101/2021.10.12.21264707

**Authors:** Manuela Di Fusco, Mary M Moran, Alejandro Cane, Daniel Curcio, Farid Khan, Deepa Malhotra, Andy Surinach, Amanda Miles, David Swerdlow, John M McLaughlin, Jennifer L Nguyen

**Author notes:** **Corresponding author** Manuela Di Fusco, Patient and Health Impact, Health Economics and Outcomes Research, Pfizer, Inc., New York, NY, USA.

## Abstract

**Objective:** To evaluate COVID-19 vaccine breakthrough infections among immunocompromised (IC) individuals.

**Methods:** Individuals vaccinated with BNT162b2 were selected from the US HealthVerity database (12/10/2020-7/8/2021). COVID-19 vaccine breakthrough infections were examined in fully vaccinated (≥14 days after 2^nd^ dose) IC individuals (IC cohort), 12 mutually exclusive IC condition groups, and a non-IC cohort. IC conditions were identified using an algorithm based on diagnosis codes and immunosuppressive (IS) medication usage.

**Results:** Of 1,277,747 individuals ≥16 years of age who received 2 BNT162b2 doses, 225,796 (17.7%) were identified as IC (median age: 58 years; 56.3% female). The most prevalent IC conditions were solid malignancy (32.0%), kidney disease (19.5%), and rheumatologic/inflammatory conditions (16.7%). Among the fully vaccinated IC and non-IC cohorts, a total of 978 breakthrough infections were observed during the study period; 124 (12.7%) resulted in hospitalization and 2 (0.2%) were inpatient deaths. IC individuals accounted for 38.2% (N=374) of all breakthrough infections, 59.7% (N=74) of all hospitalizations, and 100% (N=2) of inpatient deaths. The proportion with breakthrough infections was 3 times higher in the IC cohort compared to the non-IC cohort (N=374 [0.18%] vs. N=604 [0.06%]; unadjusted incidence rates were 0.89 and 0.34 per 100 person-years, respectively. Organ transplant recipients had the highest incidence rate; those with >1 IC condition, antimetabolite usage, primary immunodeficiencies, and hematologic malignancies also had higher incidence rates compared to the overall IC cohort. Incidence rates in older (≥65 years old) IC individuals were generally higher versus younger IC individuals (<65).

**Limitations:** This retrospective analysis relied on coding accuracy and had limited capture of COVID-19 vaccine receipt.

**Conclusions:** COVID-19 vaccine breakthrough infections are rare but are more common and severe in IC individuals. The findings from this large study support FDA authorization and CDC recommendations to offer a 3^rd^ vaccine dose to increase protection among IC individuals.

## Introduction

By the end of September 2021, the number of coronavirus disease 2019 (COVID-19) cases in the United States had reached over 43 million, with the number of deaths attributed to the illness approaching 700,000 [1]. Three vaccines have been issued Emergency Use Authorization (EUA) by the US Food and Drug Administration (FDA) and are available for active immunization to prevent COVID-19: BNT162b2 (Pfizer/BioNTech), mRNA1273 (Moderna), and Ad26.COV2.S (Janssen) [2-4]. The mRNA vaccines demonstrated 94-95% efficacy in preventing symptomatic COVID-19 illness in randomized clinical trials (RCTs) [5,6]. At the time of this study, BNT162b2 was the only vaccine with an approved Biologics License Application (BLA) and was the most widely administered vaccine in the US. Evidence accumulating under real-world conditions indicates that BNT162b2 is highly effective in protecting against SARS-CoV-2 infection, symptomatic COVID-19 illness, and COVID-19-related hospitalization and death, findings that are generally consistent with the RCT [5,7].

In the RCTs of the mRNA vaccines, those with an immunocompromised (IC) condition were largely excluded from the study populations [5,6]. Currently, there is limited information in the published literature on whether a suboptimal immune response to the COVID-19 mRNA vaccines among those with IC conditions [8-12] leads to reduced vaccine effectiveness (VE). Some recently conducted real-world studies that included subpopulations with IC conditions have found that while the COVID-19 vaccines were effective in this patient group, VE against SARS-CoV-2 infection, symptomatic illness, and COVID-19-related hospitalization was lower than observed in the general population, with VE ranging from 63% to 90% [13-21].

On August 12, 2021, the FDA amended the EUA for the mRNA COVID-19 vaccines to allow for the use of an additional dose for certain individuals with compromised immune systems [22]. The additional dose should be administered at least 28 days following the 2-dose regimen of the same mRNA vaccine. According to the FDA guidance, the IC group includes “solid organ transplant recipients or those who are diagnosed with conditions that are considered to have an equivalent level of immunocompromise” [22]. The Centers for Disease Control and Prevention (CDC) provided additional guidance and more specifically referred to individuals with moderate to severe IC conditions who are on active cancer treatment for hematologic malignancies, have undergone solid organ transplantation or a stem cell transplant and are taking immunosuppressive (IS) medications, have a moderate to severe primary immunodeficiency (e.g., DiGeorge syndrome, Wiskott-Aldrich syndrome), have advanced/untreated HIV infection, or are taking IS medications (e.g., high-dose corticosteroids) [23]. Further study is warranted to gain a better understanding of which IC conditions predispose individuals to reduced protection with COVID-19 mRNA vaccines to guide further prevention efforts and support clinical decision-making. Towards this aim, this study identified individuals with various IC conditions who had been vaccinated with the BNT162b2 vaccine and evaluated the clinical characteristics and clinical outcomes, with a primary outcome analysis of COVID-19 vaccine breakthrough infections, among those with and without an IC condition (ClinicalTrials.gov identifier: NCT05020145) [24].

## Methods

### Study design and data source

This retrospective study utilized data sets within the HealthVerity database (Philadelphia, PA) collected from December 10, 2020 through July 8, 2021. The HealthVerity database contains the largest all-payer (commercial, Medicare, and Medicaid) collection of US healthcare administrative data that links patient’s inpatient admissions, outpatient visits, laboratory visits, and pharmacy services, including COVID-19 tests and vaccinations. Electronic medical records (EMRs), including Veradigm data, are integrated with the administrative claims data from multiple sources to capture a relatively complete summary of an individual patient’s clinical history, including comorbid conditions and other risk factors that may predispose the individual to infections and complications. All data sets contained within the HealthVerity database are secured and encrypted and all patient information is deidentified. As such, the data source is compliant with the Health Insurance Portability and Accountability Act (HIPAA).

### Study population

Individuals vaccinated with BNT162b2 were selected from the HealthVerity database from December 10, 2020 through July 8, 2021 based on the Current Procedural Terminology-4 (CPT-4) code: 91300, administration codes (0001A, 0002A), and National Drug Codes (NDC: 59267100001, 59267100002, 59267100003) recorded on healthcare claims [25]. The index date was defined as the date of receipt of the 1^st^ vaccine dose. Individuals included in the study population were required to be ≥16 years of age on their index date, have continuous medical enrollment for the 12-month period prior to the index date (baseline period), and to not have been diagnosed with COVID-19 during the baseline period. A COVID-19 diagnosis was identified using International Classification of Diseases, 10^th^ revision (ICD-10) code U071. The primary outcome analysis of COVID-19 vaccine breakthrough infections focused on individuals fully vaccinated with BNT162b2. “Fully vaccinated” was defined as ≥14 days after receipt of the 2^nd^ vaccine dose, consistent with the CDC definition [26]. Individuals were followed from the date they were fully vaccinated until the end of the study period or the end of continuous medical enrollment, whichever was earlier.

### Identification of individuals with IC conditions

Individuals with an IC condition were identified using a modified algorithm originally developed and validated by Greenberg et al. [27] for use in administrative claims database studies to identify IC patients with severe sepsis. This algorithm was subsequently modified and utilized by Patel et al. [28] and Hughes et al. [29] to identify IC populations from administrative claims and EMRs, respectively. For this study, we modified the original algorithm of Greenberg et al. [27] based on expert clinical input to be specific to COVID-19; the details are provided below.

Similar to the criteria of Patel et al. [28], individuals were identified as having an IC condition if they had ≥1 hospitalization or ≥2 outpatient visits on separate dates with an ICD-10 code on a healthcare claim indicating an IC condition or usage of specific IS medications during a 12-month baseline period. The IC case definition identified 9 groups based on clinical diagnoses and 2 other groups based on usage of IS medications only, for a total of 11 groups with an IC condition (Figure 1). Individuals with >1 IC condition were also assessed, for a total of 12 mutually exclusive groups for inclusion in this study. The list of ICD-10 codes used to identify IC cases by diseases and list of IS medications are shown in Supplementary Tables 1 and 2, respectively.

**Figure 1.**
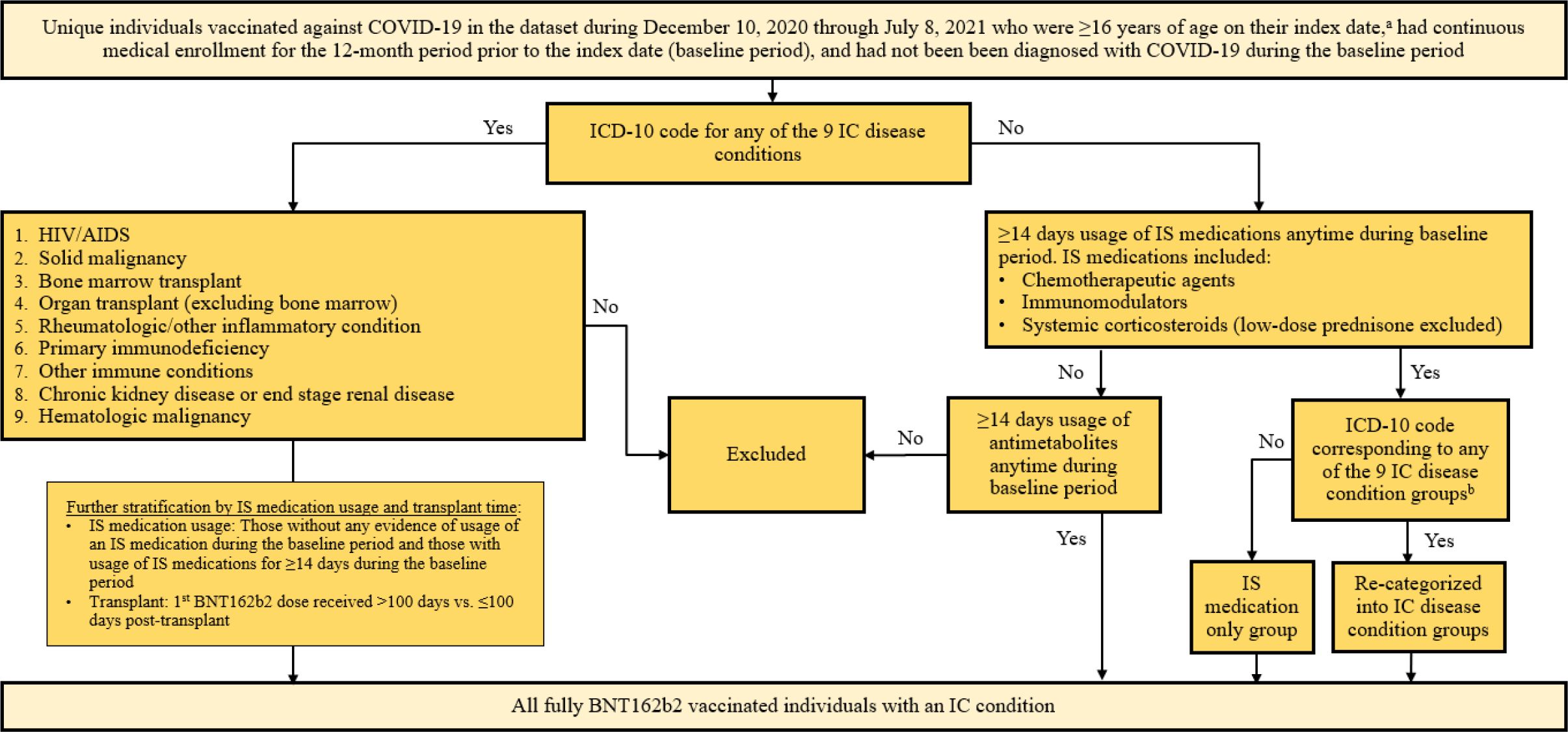
Algorithm to define IC cases. ^a^ Index date was defined as the date of receipt of the 1^st^ BNT162b2 dose. ^b^ Based on a review of ICD-10 diagnosis codes and keyword search including immune, malignancy, neoplasm, rheumatoid, and kidney, excluding encounter/screening test. AIDS: Acquired immunodeficiency syndrome; HIV: human immunodeficiency virus; IC: Immunocompromised; IS: Immunosuppressive

The disease IC condition groups included 1) symptomatic human immunodeficiency virus (HIV)/acquired immunodeficiency syndrome (AIDS); 2) solid malignancy; 3) bone marrow transplant; 4) organ transplant (excluding bone marrow transplant); 5) rheumatologic or other inflammatory condition; 6) a primary immunodeficiency; 7) other immune conditions; 8) chronic kidney disease (CKD) or end stage renal disease (ESRD); and 9) hematologic malignancy. To discern whether treatment with an IS medication may impact the occurrence of COVID-19 vaccine breakthrough infections, individuals in these 9 IC groups were further stratified by treatment status: those without any evidence of usage of an IS medication during the baseline period and those with usage of IS medications for ≥14 days during the baseline period.

Additionally, among the latter, the proportions with active treatment, defined as evidence of medication usage ≤14 days before a 1^st^ BNT162b2 dose, except for hematological malignancy, where active treatment status was defined as evidence of any IS medication (including radiotherapy) usage within 6 months before a 1^st^ BNT162b2 dose, were assessed. The IS medications included chemotherapeutic agents, immunomodulators, and systemic corticosteroids excluding low-dose (<60mg/day) prednisone; radiation therapy was included for solid malignancy. Individuals with a bone marrow or organ transplant were further stratified by those who received their 1^st^ BNT162b2 dose >100 days prior to transplant and those who received it ≤100 days prior to transplant.

The 10^th^ IC condition group included those individuals with usage of an IS medication (chemotherapeutic agent, immunomodulator, or systemic corticosteroid excluding low-dose [<60mg/day] prednisone) for ≥14 days during the baseline period, but without any ICD-10 code for the above listed IC conditions on a healthcare claim during the baseline period. Those with ≥14 days usage of antimetabolites anytime during the baseline period were excluded from this group but were placed in the 11^th^ IC condition group.

The algorithm used to define IC cases is illustrated in Figure 1. This algorithm differs from that used in Greenberg et al. [27] by the following modifications: the inclusion of individuals with a solid malignancy who had radiation therapy; the separation of individuals with a bone marrow transplant and those with an organ transplant; the inclusion of chronic inflammatory demyelinating polyneuropathy and immune thrombocytopenic purpura in the “rheumatologic or other inflammatory condition” group; the inclusion of sickle cell disease, asplenia, and psoriatic arthritis in the “other immune disorders” group; the addition of individuals with primary immunodeficiencies and those with CKD or ESRD; the reclassification of individuals with only usage of an IS medication; the removal of individuals with usage of low dose prednisone; and the inclusion of an ad-hoc group of individuals who were on antimetabolite therapies. Additionally, the following subcategories were created: the first 9 IC condition groups were stratified according to active treatment status, and organ transplant recipients were stratified according to time of receipt of the 1^st^ vaccine dose in relation to the date of transplant.

All IC condition groups were mutually exclusive. Individuals meeting the criteria for classification into a single IC condition group were categorized accordingly into IC condition groups 1-11. Any individuals meeting the criteria for classification into multiple IC condition groups were categorized into the >1 IC condition group (12^th^ IC condition group). All individuals included in the study population were then grouped into study cohorts consisting of those with an IC condition (IC cohort) and those without an IC condition (non-IC cohort).

### Demographic and clinical characteristics

Demographic characteristics, including age, sex, US census region of residence, and payer type, were extracted from the data source for all individuals who had received 2 doses of the BNT162b2 vaccine. Medical conditions that determine whether or not an individual may be at risk of severe COVID-19, according to the Advisory Committee on Immunization Practices (ACIP) of the CDC and FDA Sentinel Master Protocol, were also examined [30,31]. Additionally, the vaccine administration setting and characteristics indicating healthcare prevention seeking behavior (i.e., had ≥1 telehealth or telephone visit, or ≥1 COVID-19 laboratory test, or receipt of the influenza vaccine) anytime during the baseline period were explored; see Supplementary Table 3 for the CPT-4 codes used. Among the individuals identified with an IC condition, the prevalence of each IC condition and IS medication treatment patterns were also evaluated.

### Vaccine utilization

The proportions of study cohorts who received their 1^st^ BNT162b2 dose stratified by month were calculated. Additionally, the proportions fully vaccinated with ≥14 days of follow-up after their 2^nd^ vaccine dose among those who received a 1^st^ vaccine dose were determined. The duration in days between 1^st^ and 2^nd^ vaccine doses was also reported.

### Study outcome measures

The primary outcome was the incidence of COVID-19 vaccine breakthrough infections, defined as the diagnosis of COVID-19 identified by ≥1 ICD-10 code of U071 on a healthcare claim ≥14 days following a 2^nd^ BNT162b2 dose. The proportion of patients with COVID-19 vaccine breakthrough infections in the IC cohort, the 12 IC condition subgroups, the non-IC cohort, and total study population were reported. Incidence rates were calculated as the number of breakthrough infections divided by the time at risk per 100 person-years and were reported with Poisson 95% confidence intervals (CIs).

The clinical presentation and severity, demographic characteristics, and time to infection for breakthrough infection cases were evaluated. Healthcare resource utilization among breakthrough infection cases, including any outpatient encounters (emergency department [ED] visit, outpatient hospital visit, other outpatient encounter) and hospital admissions, with a breakdown of invasive mechanical ventilation (IMV) usage and intensive care unit (ICU) admission, was also examined. ICU admission and IMV usage were identified using revenue codes, Healthcare Common Procedure Coding System (HCPCS) codes, and/or CPT-4 codes. IMV included extracorporeal membrane oxygenation (ECMO).

### Statistical analyses

All study outcome measurements were summarized with descriptive statistics. Counts and percentages were reported for categorical variables. Means, standard deviations, medians, and first (Q1) and third (Q3) quartiles were reported for continuous variables. Statistical analyses were performed using SAS statistical software, version 9.4 (SAS Institute Inc., Cary, North Carolina).

### Subgroup analyses

Subgroup analyses were conducted by age (<65 and ≥65 years), by active treatment status, and for organ transplant recipients. The active treatment status analyses explored the number of COVID-19 vaccine breakthrough infections that occurred in the 4 CDC IC groups for which an additional dose is currently recommended based on active treatment status (i.e., organ transplant, hematological malignancy, IS medication usage, antimetabolite usage) [23], with and without the active treatment requirement. Active treatment was defined as evidence of medication usage ≤14 days before a 1^st^ BNT162b2 dose, except for hematological malignancy, where active treatment status was defined as evidence of any IS medication (including radiotherapy) usage within 6 months before a 1^st^ BNT162b2 dose.

### Ethics statement

The study followed the Strengthening the Reporting of Observational Studies in Epidemiology (STROBE) reporting guideline [32]. This study was deemed exempt from Institutional Review Board (IRB) review pursuant to the terms of the US Department of Health and Human Service’s Policy for Protection of Human Research Subjects at 45 C.F.R. 46.104(d); category 4 exemption (Sterling IRB, Boston, Maine waived ethical approval for this work). This study was registered at ClinicalTrials.gov (NCT05020145) [24].

## Results

### Study population

A total of 21,725,804 individuals vaccinated against COVID-19 were identified from December 10, 2020 through July 8, 2021 in the HealthVerity database; 4,375,051 individuals met the study inclusion criteria on index dates. Among this study population, 778,106 (17.8%) met the criteria of having ≥1 IC condition and the remaining 3,596,945 (82.2%) were categorized as non-IC. A total of 1,277,747 individuals received 2 doses of BNT162b2, of which 225,796 (17.7%) were grouped into the IC cohort and 1,051,951 (82.3%) were placed into the non-IC cohort. A total of 1,176,907 (92.1%) individuals had ≥14 days of follow-up after the 2^nd^ BNT162b2 dose; approximately 94% (N=212,945) and 92% (N=963,962) in the IC cohort and non-IC cohort, respectively. The population attrition for meeting study selection criteria is shown in Figure 2.

**Figure 2.**
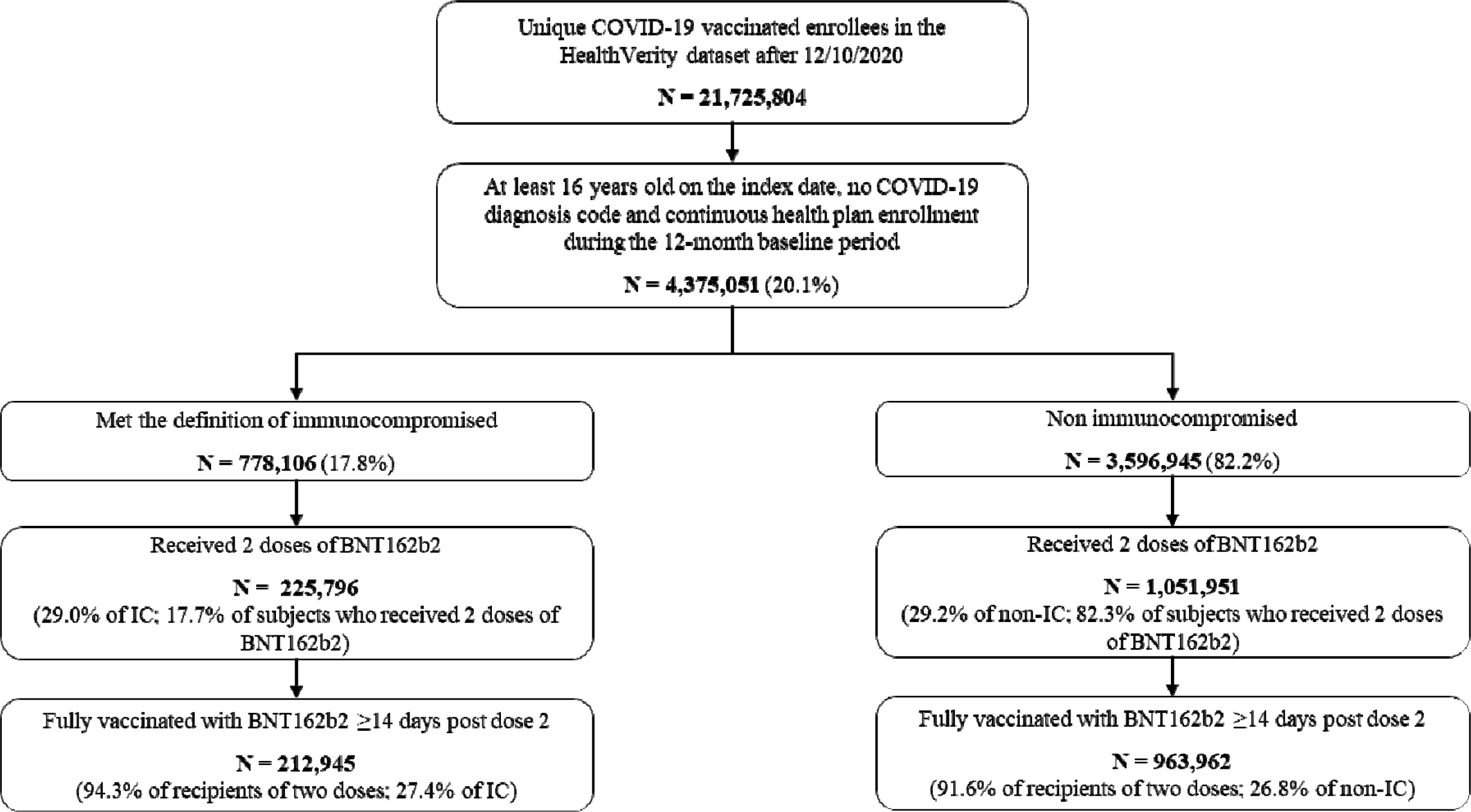
Study selection criteria. IC: Immunocompromised

### Demographic and clinical characteristics

The characteristics of the IC cohort, non-IC cohort, and overall study population that received 2 BNT162b2 doses are shown in Table 1. The median age of individuals in the IC-cohort was 58 years, with approximately one-half (48.3%) between 50 and 64 years of age and 23.0% ≥65 years of age. In the IC cohort, 56.3% were female and 74.5% had commercial insurance coverage. The median age of individuals in the non-IC cohort was 45 years, with one-third (33.3%) between 50 and 64 years of age and 9.3% ≥65 years of age. In the non-IC cohort, 53.1% were female and 80.2% had commercial insurance coverage. Nearly two-thirds (62.5%) of individuals in the IC cohort had ≥1 condition indicating they were at risk of severe COVID-19, compared with only 19.3% of individuals in the non-IC cohort. Those in the IC cohort showed greater healthcare prevention seeking behavior (i.e., had ≥1 telehealth or telephone visit, or ≥1 COVID-19 laboratory test, or receipt of the influenza vaccine) than those in the non-IC cohort during the baseline period prior to having received the 1^st^ BNT162b2 dose.

**Table 1.**
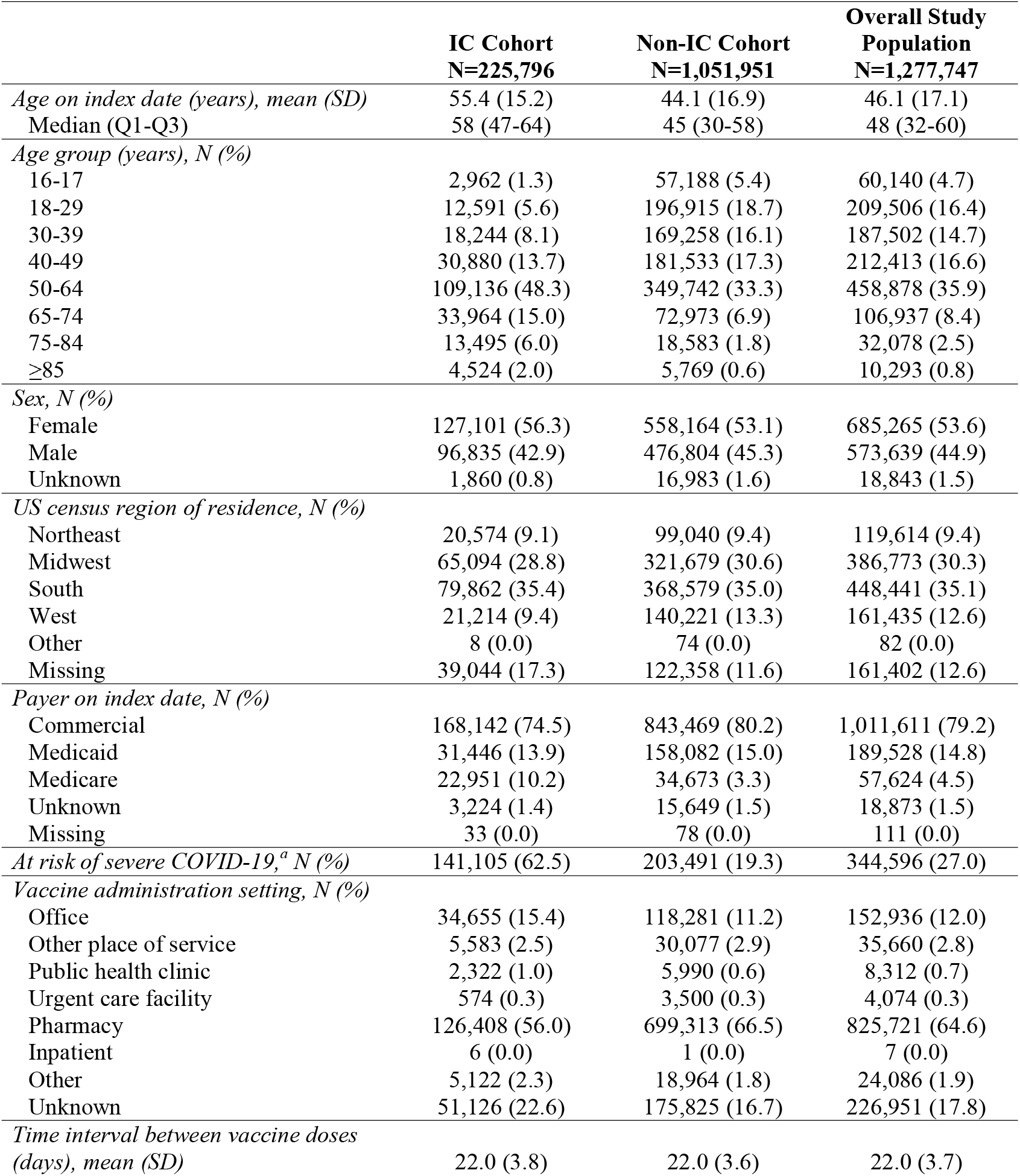
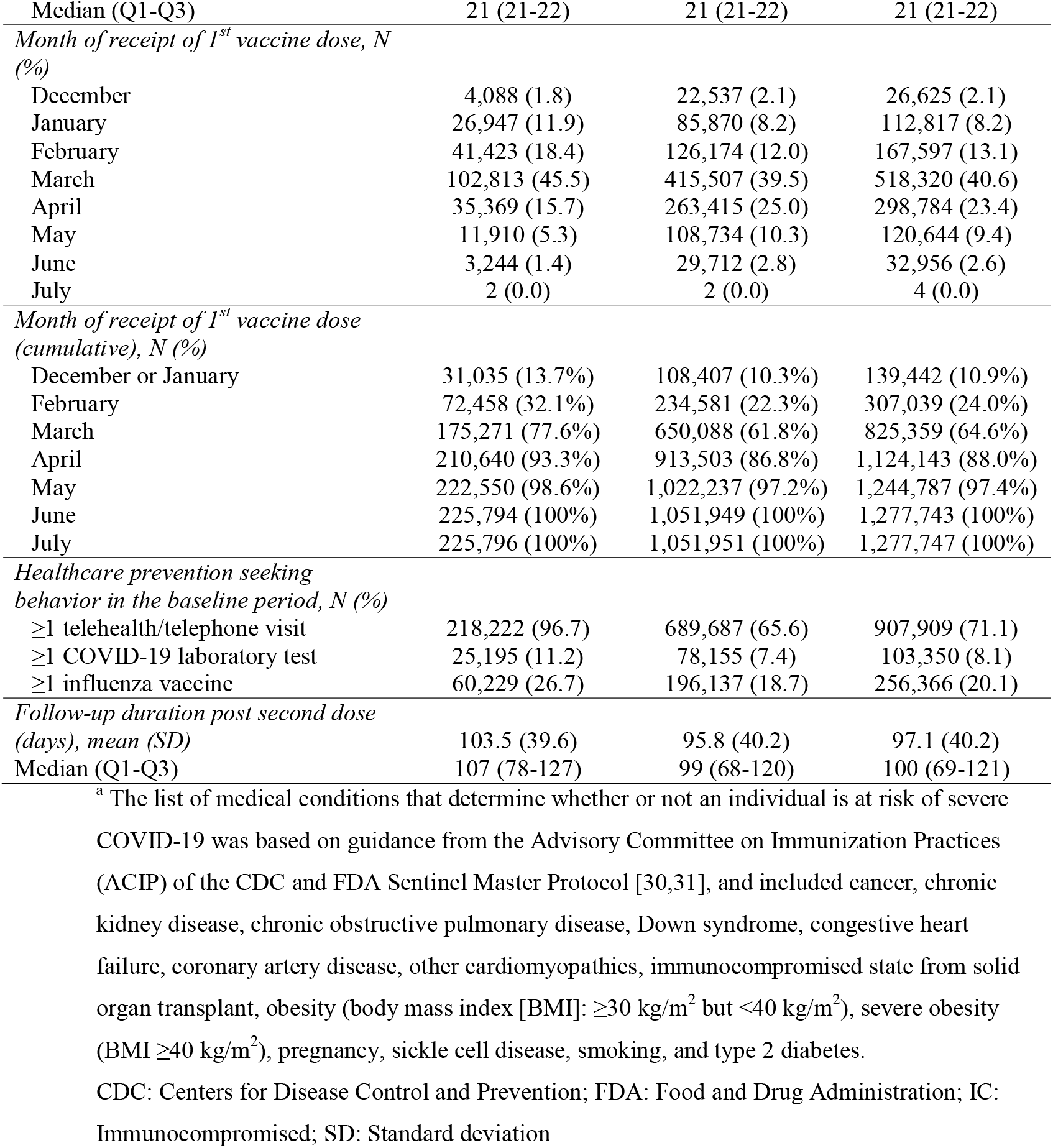
Characteristics of the IC and non-IC cohorts and overall study population that received 2 BNT162b2 doses.

### Prevalence of IC conditions and IS medication treatment status

Among the IC cohort that received 2 BNT162b2 doses, the most prevalent IC conditions were solid malignancy (32.0%), kidney disease (19.5%), and rheumatologic/inflammatory conditions (16.7%); 16.9% had >1 IC condition (Supplementary Table 4). Approximately one-fifth (21.2%) of individuals in the IC cohort used IS medication for ≥14 days anytime during the baseline period, with 4.2% having active treatment (i.e., ≤14 days prior to receipt of their 1^st^ BNT162b2 dose; Supplementary Table 4). The IC condition groups with the highest prevalence of active treatment with IS medications were organ transplant recipients (20.0%), and those with rheumatologic/other inflammatory conditions (7.7%), >1 IC condition (6.5%), and hematologic malignancy (4.2%). Among those with IS medication usage and antimetabolite usage (and no other IC condition), the prevalence of active treatment was 11.9% and 29.0%, respectively (Supplementary Table 4).

### Vaccine utilization

The cumulative proportions of individuals who received their 1^st^ BNT162b2 dose by month are shown in Table 1. More than three-quarters (77.6%) of the IC cohort had received their 1^st^ dose by the end of March, versus 61.8% in the non-IC cohort. The proportions fully vaccinated with BNT162b2 (i.e., with ≥14 days of follow-up after their 2^nd^ dose) were similar among the IC cohort (27.4% of total IC population) and the non-IC cohort (26.8% of total non-IC population) (Figure 2). Among those who were fully vaccinated across both the IC and non-IC cohorts, the mean duration between 1^st^ and 2^nd^ doses was 22 days and median duration was 21 days (Q1-Q3: 21-22 days) (Table 1). The majority of individuals in the IC (56.0%) and non-IC (66.5%) cohorts received the 1^st^ BNT162b2 dose at the pharmacy as opposed to a physician’s office or other setting (Table 1).

### Incidence and clinical presentation of COVID-19 vaccine breakthrough infections

During the study period (December 10, 2020-July 8, 2021), a total of 978 COVID-19 vaccine breakthrough infections were observed among those who were fully vaccinated with BNT162b2 (Table 2). Among those with breakthrough infections, median age was 61 years in the IC cohort and 51 years in the non-IC cohort; 58.0% and 60.0% were female, respectively (Table 3). The proportion with breakthrough infections was 3 times higher in the IC cohort compared to the non-IC cohort (N=374 [0.18%] vs. N=604 [0.06%]) (Table 2). Unadjusted incidence rates were 0.89 and 0.34, respectively, per 100 person-years, for a 2.6-fold increased rate in IC versus non-IC (Table 2).

**Table 2.** Incidence rates of COVID-19 vaccine breakthrough infections per 100 person-years among individuals fully vaccinated^a^ with BNT162b2, by IC condition and age <65 and ≥65 years.

| **Study population** | **N** | **Breakthrough infections** | **Proportion** | **Person-years<sup>b</sup>** | **IR per 100 person-years** | **Lower 95% CI** | **Upper 95% CI** |
| --- | --- | --- | --- | --- | --- | --- | --- |
| *Non-IC cohort* | 963,962 | 604 | 0.06% | 175101.5 | 0.34 | 0.32 | 0.37 |
| *IC cohort* | 212,945 | 374 | 0.18% | 42069.2 | 0.89 | 0.80 | 0.98 |
| *12 IC cohorts* |  |  |  |  |  |  |  |
| HIV/AIDS | 2103 | 3 | 0.14% | 424.4 | 0.71 | 0.15 | 2.07 |
| Solid malignancy | 68,545 | 77 | 0.11% | 13780.2 | 0.56 | 0.44 | 0.70 |
| Bone marrow transplant | 12 | 0 | 0.00% | 2.6 | 0.00 | - | - |
| Organ transplant (excluding bone marrow) | 675 | 5 | 0.74% | 136.6 | 3.66 | 1.19 | 8.54 |
| Rheumatologic/other inflammatory condition | 35,475 | 57 | 0.16% | 6980.7 | 0.82 | 0.62 | 1.06 |
| Primary immunodeficiency | 3190 | 7 | 0.22% | 619.7 | 1.13 | 0.45 | 2.33 |
| Other immune condition<sup>c</sup> | 3684 | 2 | 0.05% | 729.5 | 0.27 | 0.03 | 0.99 |
| CKD or ESRD | 41,597 | 77 | 0.19% | 8084.6 | 0.95 | 0.75 | 1.19 |
| Hematological malignancy | 1814 | 4 | 0.22% | 366.4 | 1.09 | 0.30 | 2.80 |
| IS medication usage ≥14 days in baseline period | 19,074 | 17 | 0.09% | 3563.1 | 0.48 | 0.28 | 0.76 |
| Antimetabolite usage ≥14 days in baseline period | 659 | 2 | 0.30% | 135.1 | 1.48 | 0.18 | 5.35 |
| >1 IC condition | 36,117 | 123 | 0.34% | 7246.5 | 1.70 | 1.41 | 2.03 |
| **<65 years of age** | **N** | **Breakthrough infections** | **Proportion** | **Person-years<sup>b</sup>** | **IR per 100 person-years** | **Lower 95% CI** | **Upper 95% CI** |
| *Non-IC cohort* | 871,022 | 485 | 0.06% | 156666.6 | 0.31 | 0.28 | 0.34 |
| *IC cohort* | 163,235 | 228 | 0.14% | 32321.4 | 0.71 | 0.62 | 0.80 |
| *12 IC cohorts* |  |  |  |  |  |  |  |
| HIV/AIDS | 2000 | 3 | 0.15% | 400.5 | 0.75 | 0.15 | 2.19 |
| Solid malignancy | 52,900 | 46 | 0.09% | 10581.7 | 0.43 | 0.32 | 0.58 |
| Bone marrow transplant | 11 | 0 | 0.00% | 2.2 | 0.00 | - | - |
| Organ transplant (excluding bone marrow) | 538 | 4 | 0.74% | 109.9 | 3.64 | 0.99 | 9.31 |
| Rheumatologic/other inflammatory condition | 29,382 | 39 | 0.13% | 5800.0 | 0.67 | 0.48 | 0.92 |
| Primary immunodeficiency | 2729 | 7 | 0.26% | 523.7 | 1.34 | 0.54 | 2.75 |
| Other immune condition<sup>c</sup> | 3269 | 1 | 0.03% | 644.8 | 0.16 | 0.00 | 0.86 |
| CKD or ESRD | 29,760 | 39 | 0.13% | 5863.2 | 0.67 | 0.47 | 0.91 |
| Hematological malignancy | 1315 | 2 | 0.15% | 271.1 | 0.74 | 0.09 | 2.67 |
| IS medication usage $\geq 14$ days in baseline period | 15,950 | 15 | 0.09% | 2947.6 | 0.51 | 0.28 | 0.84 |
| Antimetabolite usage $\geq 14$ days in baseline period | 552 | 2 | 0.36% | 112.8 | 1.77 | 0.21 | 6.40 |
| >1 IC condition | 24,829 | 70 | 0.28% | 5063.8 | 1.38 | 1.08 | 1.75 |

| $\geq 65$ years of age | N | Breakthrough infections | Proportion | Person-years<sup>b</sup> | IR per 100 person-years | Lower 95% CI | Upper 95% CI |
| --- | --- | --- | --- | --- | --- | --- | --- |
| *Non-IC cohort* | 92,940 | 119 | 0.13% | 18434.9 | 0.65 | 0.53 | 0.77 |
| *IC cohort* | 49,710 | 146 | 0.29% | 9747.8 | 1.50 | 1.26 | 1.76 |
| *12 IC cohorts* |  |  |  |  |  |  |  |
| HIV/AIDS | 103 | 0 | 0.00% | 23.9 | 0.00 | - | - |
| Solid malignancy | 15,645 | 31 | 0.20% | 3198.4 | 0.97 | 0.66 | 1.38 |
| Bone marrow transplant | 1 | 0 | 0.00% | 0.3 | 0.00 | - | - |
| Organ transplant (excluding bone marrow) | 137 | 1 | 0.73% | 26.7 | 3.75 | 0.09 | 20.90 |
| Rheumatologic/other inflammatory condition | 6093 | 18 | 0.30% | 1180.6 | 1.52 | 0.90 | 2.41 |
| Primary immunodeficiency | 461 | 0 | 0.00% | 96.0 | 0.00 | - | - |
| Other immune condition<sup>c</sup> | 415 | 1 | 0.24% | 84.7 | 1.18 | 0.03 | 6.58 |
| CKD or ESRD | 11,837 | 38 | 0.32% | 2221.4 | 1.71 | 1.21 | 2.35 |
| Hematological malignancy | 499 | 2 | 0.40% | 95.3 | 2.10 | 0.25 | 7.58 |
| IS medication usage $\geq 14$ days in baseline period | 3124 | 2 | 0.06% | 615.5 | 0.32 | 0.04 | 1.17 |
| Antimetabolite usage $\geq 14$ days in baseline period | 107 | 0 | 0.00% | 22.2 | 0.00 | - | - |
| >1 IC condition | 11,288 | 53 | 0.47% | 2182.7 | 2.43 | 1.82 | 3.18 |

<sup>a</sup> Fully vaccinated was defined as $\geq 14$ days after receipt of the 2<sup>nd</sup> BNT162b2 dose.
<sup>b</sup> Person time was calculated starting from 14 days after the receipt of the 2<sup>nd</sup> dose of BNT162b2 until a COVID-19 vaccine breakthrough infection case or end of continuous enrollment, whichever occurred earlier.
<sup>c</sup> Other immune conditions are described in Supplementary Table 1.
AIDS: Acquired immunodeficiency syndrome; CI: Confidence interval; CKD: Chronic kidney disease; ESRD: End stage renal disease; HIV: human immunodeficiency virus; IC: Immunocompromised; IR: Incidence rate; IS: Immunosuppressive

**Table 3.**
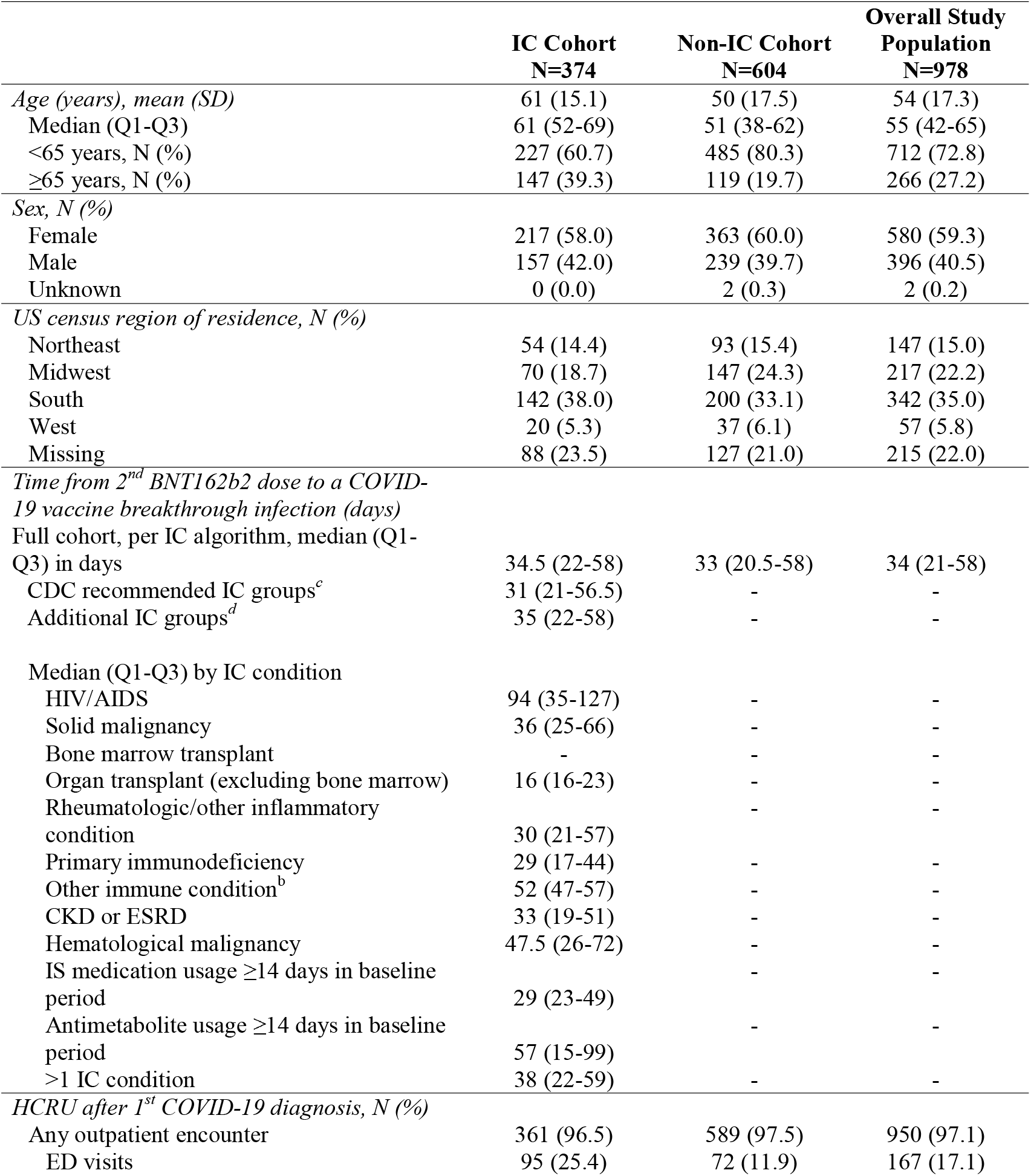
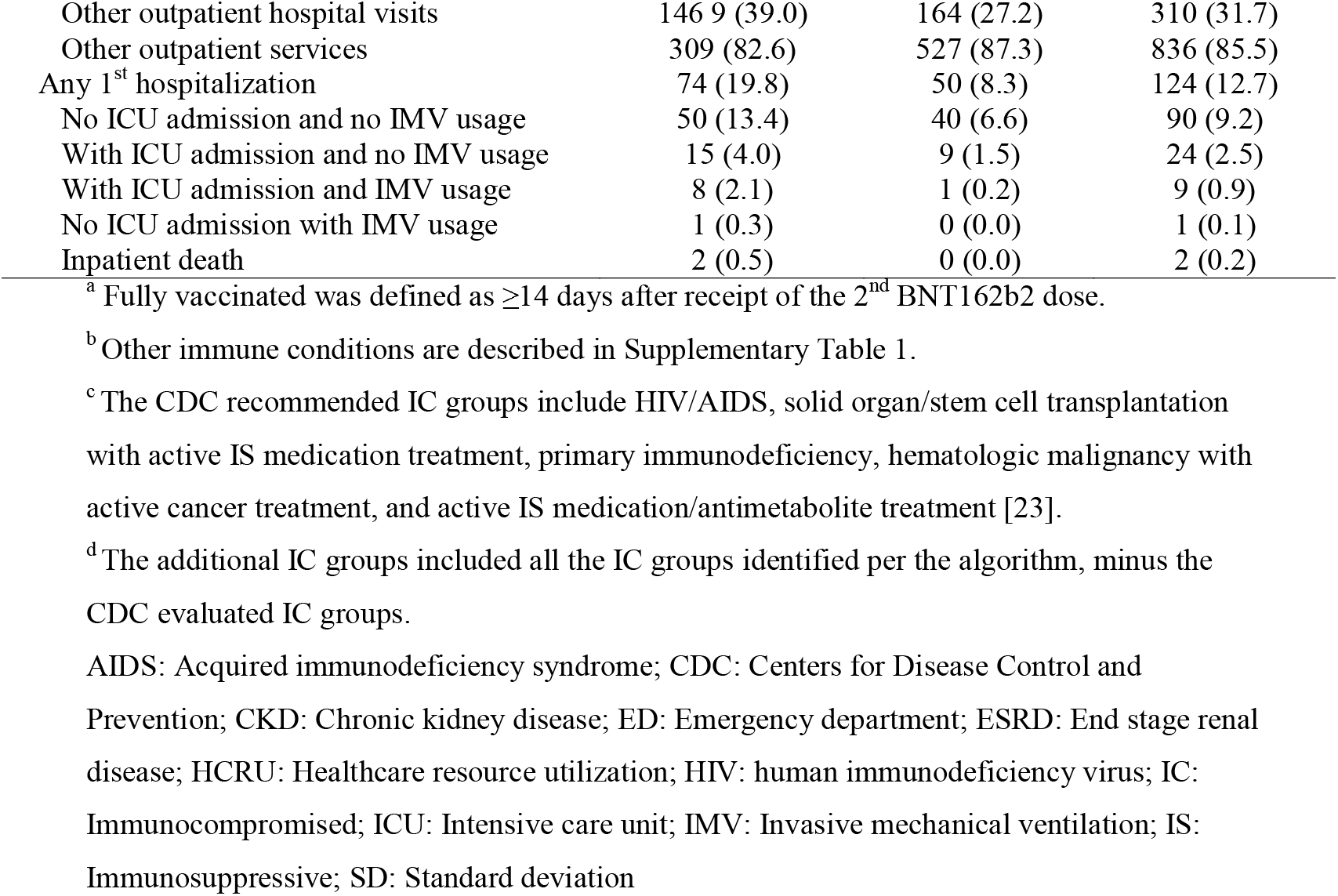
Characteristics of fully vaccinated^a^ individuals with COVID-19 vaccine breakthrough infections.

|  | **IC Cohort**<br>**N=374** | **Non-IC Cohort**<br>**N=604** | **Overall Study**<br>**Population**<br>**N=978** |
| --- | --- | --- | --- |
| *Age (years), mean (SD)* | 61 (15.1) | 50 (17.5) | 54 (17.3) |
| Median (Q1-Q3) | 61 (52-69) | 51 (38-62) | 55 (42-65) |
| <65 years, N (%) | 227 (60.7) | 485 (80.3) | 712 (72.8) |
| ≥65 years, N (%) | 147 (39.3) | 119 (19.7) | 266 (27.2) |
| *Sex, N (%)* |  |  |  |
| Female | 217 (58.0) | 363 (60.0) | 580 (59.3) |
| Male | 157 (42.0) | 239 (39.7) | 396 (40.5) |
| Unknown | 0 (0.0) | 2 (0.3) | 2 (0.2) |
| *US census region of residence, N (%)* |  |  |  |
| Northeast | 54 (14.4) | 93 (15.4) | 147 (15.0) |
| Midwest | 70 (18.7) | 147 (24.3) | 217 (22.2) |
| South | 142 (38.0) | 200 (33.1) | 342 (35.0) |
| West | 20 (5.3) | 37 (6.1) | 57 (5.8) |
| Missing | 88 (23.5) | 127 (21.0) | 215 (22.0) |
| *Time from 2<sup>nd</sup> BNT162b2 dose to a COVID-19 vaccine breakthrough infection (days)* |  |  |  |
| Full cohort, per IC algorithm, median (Q1-Q3) in days | 34.5 (22-58) | 33 (20.5-58) | 34 (21-58) |
| CDC recommended IC groups<sup>c</sup> | 31 (21-56.5) | - | - |
| Additional IC groups<sup>d</sup> | 35 (22-58) | - | - |
| *Median (Q1-Q3) by IC condition* |  |  |  |
| HIV/AIDS | 94 (35-127) | - | - |
| Solid malignancy | 36 (25-66) | - | - |
| Bone marrow transplant | - | - | - |
| Organ transplant (excluding bone marrow) | 16 (16-23) | - | - |
| Rheumatologic/other inflammatory condition | 30 (21-57) | - | - |
| Primary immunodeficiency | 29 (17-44) | - | - |
| Other immune condition<sup>b</sup> | 52 (47-57) | - | - |
| CKD or ESRD | 33 (19-51) | - | - |
| Hematological malignancy | 47.5 (26-72) | - | - |
| IS medication usage ≥14 days in baseline period | 29 (23-49) | - | - |
| Antimetabolite usage ≥14 days in baseline period | 57 (15-99) | - | - |
| >1 IC condition | 38 (22-59) | - | - |
| *HCRU after 1<sup>st</sup> COVID-19 diagnosis, N (%)* |  |  |  |
| Any outpatient encounter | 361 (96.5) | 589 (97.5) | 950 (97.1) |
| ED visits | 95 (25.4) | 72 (11.9) | 167 (17.1) |
| Other outpatient hospital visits | 146 9 (39.0) | 164 (27.2) | 310 (31.7) |
| Other outpatient services | 309 (82.6) | 527 (87.3) | 836 (85.5) |
| Any 1<sup>st</sup> hospitalization | 74 (19.8) | 50 (8.3) | 124 (12.7) |
| No ICU admission and no IMV usage | 50 (13.4) | 40 (6.6) | 90 (9.2) |
| With ICU admission and no IMV usage | 15 (4.0) | 9 (1.5) | 24 (2.5) |
| With ICU admission and IMV usage | 8 (2.1) | 1 (0.2) | 9 (0.9) |
| No ICU admission with IMV usage | 1 (0.3) | 0 (0.0) | 1 (0.1) |
| Inpatient death | 2 (0.5) | 0 (0.0) | 2 (0.2) |

<sup>a</sup> Fully vaccinated was defined as $\geq 14$ days after receipt of the 2<sup>nd</sup> BNT162b2 dose.
<sup>b</sup> Other immune conditions are described in Supplementary Table 1.
<sup>c</sup> The CDC recommended IC groups include HIV/AIDS, solid organ/stem cell transplantation with active IS medication treatment, primary immunodeficiency, hematologic malignancy with active cancer treatment, and active IS medication/antimetabolite treatment [23].
<sup>d</sup> The additional IC groups included all the IC groups identified per the algorithm, minus the CDC evaluated IC groups.
AIDS: Acquired immunodeficiency syndrome; CDC: Centers for Disease Control and Prevention; CKD: Chronic kidney disease; ED: Emergency department; ESRD: End stage renal disease; HCRU: Healthcare resource utilization; HIV: human immunodeficiency virus; IC: Immunocompromised; ICU: Intensive care unit; IMV: Invasive mechanical ventilation; IS: Immunosuppressive; SD: Standard deviation

The incidence rate of breakthrough infections was highest among organ transplant recipients (excluding bone marrow transplant recipients) at 3.66 per 100 person-years. Certain IC conditions had incidence rates that were higher than the rate for the overall IC cohort, specifically those with >1 IC condition (1.70 per 100 person-years), those with ≥14 days usage of antimetabolites anytime during the baseline period (1.48 per 100 person-years), those with a primary immunodeficiency (1.13 per 100 person-years), those with a hematologic malignancy (1.09 per 100 person-years) and those with CKD or ESRD (0.95 per 100 person-years) (Table 2).

A total of 950 (97.1%) COVID-19 vaccine breakthrough infections resulted in an outpatient encounter, 124 (12.7%) in hospitalization, and 2 (0.2%) in inpatient deaths (Table 3). Among those with breakthrough infections in the IC cohort (N=374), 361 (96.5%) had an outpatient encounter and 74 (19.8%) were hospitalized; during hospitalization, 9 (2.4%) required IMV, 23 (6.1%) were admitted to the ICU, and 2 (0.5%) died (Table 3). While individuals in the IC cohort only represented approximately 18% of the fully vaccinated population, they accounted for 38.2% of all breakthrough infections, 59.7% of all hospitalizations, and 100% of inpatient deaths.

Among those with breakthrough infections in the non-IC cohort (N=604), 589 (97.5%) had an outpatient encounter and 50 (8.3%) were hospitalized; during hospitalization, 1 (0.2%) required IMV, 10 (1.7%) were admitted to the ICU, and none died (Table 3).

### Time to COVID-19 vaccine breakthrough infection

The median duration post-2^nd^ BNT162b2 dose to a COVID-19 diagnosis was 34.5 days (Q1-Q3: 22-58 days) for the IC cohort and 33 days (Q1-Q3: 20.5-58 days) for the non-IC cohort (Table 3). The median duration for the CDC recommended IC groups (N=32) [23] was 31 days (Q1-Q3: 21-56.5 days), slightly shorter than the overall IC cohort and the non-IC cohort. The median duration for the additional IC groups was 35 days (Q1-Q3: 22-58 days). Organ transplant recipients had the shortest time to infection post-2^nd^ BNT162b2 dose, a median of 16 days (Q1-Q3: 16-23 days). Those with HIV/AIDS and antimetabolite usage had the longest times to infection, a median of 94 days (Q1-Q3: 35-127 days) and 57 days (Q1-Q3: 15-99 days), respectively (Table 3).

### Subgroup analyses: Age groups

Among the fully vaccinated IC cohort, a total of 163,235 individuals were <65 years of age on their index date, representing 76.7% of the population in this cohort (Table 2). In the subgroup <65 years of age, 228 COVID-19 vaccine breakthrough infections were observed, representing 61.0% of all breakthrough infections in the IC cohort; the incidence rate for this subgroup was 0.71 per 100 person-years. For IC individuals ≥65 years of age (N=49,710), the incidence rate was 1.50 per 100 person-years. Among this older subgroup, incidence rates were higher for those with solid malignancy, rheumatologic/inflammatory conditions, CKD or ESRD, other immune conditions, hematologic malignancy, and >1 IC condition than among those <65 years of age with the same corresponding IC conditions.

Among the fully vaccinated non-IC cohort, 90.4% (N=871,022) were <65 years of age on their index date (Table 2). They accounted for 80.3% (N=485) of all COVID-19 vaccine breakthrough infections in the non-IC cohort; the incidence rate for this subgroup was 0.31 per 100 person-years. For those ≥65 years of age (N=92,940), the incidence rate was 0.65 per 100 person-years (Table 2).

### Subgroup analyses: Organ transplant recipients

Among organ transplant recipients (excluding bone marrow transplant recipients) who received 2 BNT162b2 doses (N=710), median age was 56 years and 43.0% were female (Supplementary Table 5). Over two-thirds (71.4%) had their transplant ≤100 days prior to receipt of their 1^st^ BNT162b2 dose (Supplementary Table 5). Regardless of age, fully vaccinated organ transplant recipients (N=675) had the highest incidence rate of COVID-19 vaccine breakthrough infections (3.64-3.75 per 100 person-years) (Table 2) and the shortest time to infection (Table 3) of the IC condition groups. The median time to infection was shorter among organ transplant recipients than among the overall IC cohort (16 vs. 34.5 days, Table 3).

### Subgroup analyses: Active treatment status

When we limited the primary outcome analysis to the 4 IC groups for which an additional vaccine dose is recommended by the CDC according to active treatment status, 6 COVID-19 vaccine breakthrough infections were observed: 1 in an organ transplant recipient, 2 in patients with hematological malignancy, and 3 in patients with evidence of IS medication usage (excluding antimetabolites) (Figure 3). When relaxing the active treatment status requirement in these 4 recommended IC groups, we observed 22 additional breakthrough infections: 4 more among organ transplant recipients, 2 more among those with hematological malignancy, 14 more among those with evidence of IS medication usage (not antimetabolites), and 2 among those with evidence of antimetabolite usage (Figure 3).

**Figure 3.**
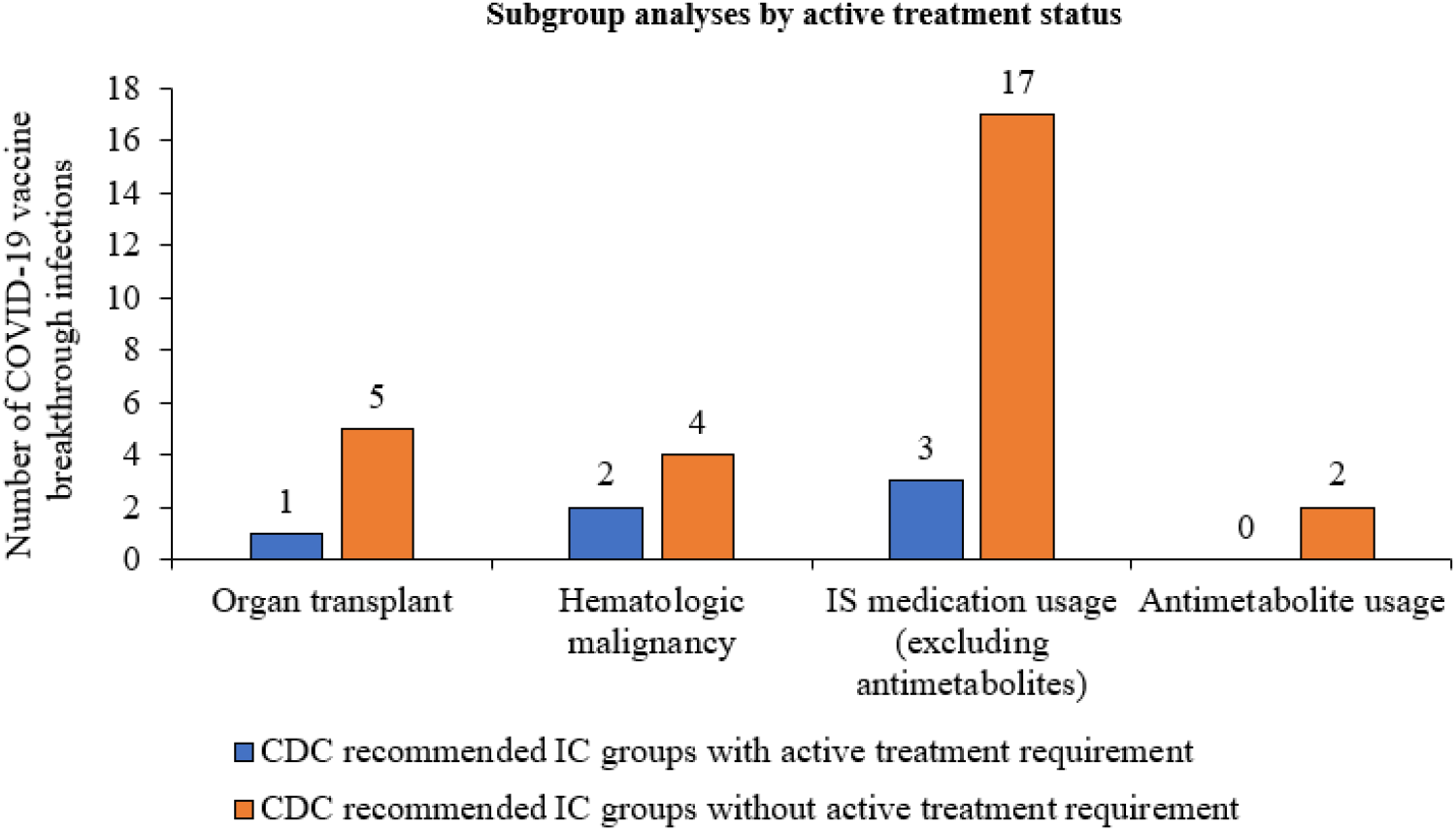
COVID-19 vaccine breakthrough infections: subgroup analysis of CDC recommended IC groups by active treatment status. Active treatment was defined as evidence of medication usage ≤14 days before a 1^st^ BNT162b2 dose, except for hematological malignancy, where active treatment status was defined as evidence of any IS medication (including radiotherapy) usage within 6 months before a 1^st^ BNT162b2 dose. CDC: Centers for Disease Control and Prevention; IC: Immunocompromised; IS: Immunosuppressive

## Discussion

This real-world US observational study included nearly 1.2 million people fully vaccinated with BNT162b2, of which over 212,000 (18%) had an IC condition as defined by the IC case condition algorithm used in this study. From December 10, 2020 up to July 8^th^, 2021, among the total fully vaccinated population, the number of COVID-19 vaccine breakthrough infections observed was low (N=978; 0.08%). Almost 40% of all breakthrough infections, approximately 60% of those breakthrough infections resulting in hospitalization, and 100% of those resulting in inpatient death, occurred in the IC cohort. Among the total population of the IC cohort, the incidence rate of breakthrough infections was 2.6 times higher than in the non-IC cohort (0.89 vs. 0.34 per 100 person-years). Organ transplant recipients (excluding bone marrow transplant recipients) had the highest incidence rate of breakthrough infections (3.66 per 100 person-years). Incidence rates were also higher than the rate for the overall IC cohort among those with >1 IC condition, those with ≥14 days usage of antimetabolites anytime during the baseline period, those with a primary immunodeficiency, those with a hematologic malignancy, and those with kidney disease. These findings support FDA authorization and CDC recommendations to offer a 3^rd^ vaccine dose to increase protection among IC individuals [22,23]. Additionally, the study findings generate novel insights into post-vaccination outcomes for the IC population using a broader spectrum of conditions, which may guide further risk-mitigation strategies.

While some COVID-19 vaccine breakthrough infections among those who are fully vaccinated against COVID-19 are expected, the findings of this study show that they are rare and less likely to result in hospitalization or death in those without an IC condition. However, further research is necessary to continue monitoring the rates of breakthrough infections in the general population, especially with waning duration of protection and emerging SARS-CoV-2 variants.

Regarding the IC population in this study, the broader algorithm we used to define IC cases resulted in a IC prevalence that was approximately 15% higher than the CDC’s estimate of ∼3% among US adults, which specifically includes those with HIV/AIDS, solid organ/stem cell transplantation with active IS medication treatment, primary immunodeficiency, hematologic malignancy with active cancer treatment, and active IS medication/antimetabolite treatment [23]. When we applied the CDC definition of IC recommended groups [23] to the population in the current study, we identified 2.2% as IC, which is relatively consistent to the CDC’s estimate of ∼3%. With the more comprehensive IC case algorithm used herein, we were able to identify more broadly individuals who were IC and may be at higher risk for breakthrough infections. Furthermore, we observed that breakthrough infections occurred in the CDC recommended IC groups regardless of active IS medication treatment status. It should be noted that the IC population identified in this study may differ from IC patients included in other studies in terms of demographics, clinical characteristics, and other factors, such as timing of vaccine receipt. Future studies may be needed to further compare breakthrough infection rates and time to infection among IC populations defined according to the CDC definition versus those defined by other algorithms.

The findings of this study complement and expand on those of other recent real-world studies that have indicated reduced COVID-19 vaccine protection for some IC patient groups [13-21]. In a US study of hospitalized patients, Tenforde et al. found that of the 45 hospitalized COVID-19 cases, 44% were IC; mRNA VE against COVID-19 hospitalization was estimated at 63% for those who were IC and 91% among those who were not [13]. A study conducted in Israel found that IC patients comprised 40% of the fully BNT162b2 vaccinated hospitalized patients with COVID-19 illness (N=152) and 47% of those with a poor outcome (i.e., IMV usage or death) [14]. In another Israeli study, BNT162b2 VE against SARS-CoV-2 infection was reduced by 33% in those who were IC compared to the 95% VE in the general population [15]. In a study conducted in the UK, Whitaker et al. reported a VE of 73% (after 2 doses of BNT162b2) against acute symptomatic/critical illness among those who were IC compared to 93% VE in the overall study population [16]. In another study from Israel, Chodick et al. reported a BNT162b2 VE of 71% against preventing SARS-CoV-2 infection in the IC (N=25,459) and 90% in the overall study population [17].

Other studies have reported somewhat higher VE in those who are IC [18-20]. Dagan et al. (also from a study population in Israel) reported a BNT162b2 VE of 90% against SARS-CoV-2 infection, 84% against symptomatic infection and 100% VE against hospitalization and severe disease in those who were IC (N=1,674); among those with chronic kidney disease (N=8,212), VE against symptomatic SARS-CoV-2 infection, hospitalization, and severe disease were 80%, 76%, and 74%, respectively [18,19]. In a US Veteran’s Affairs study, mRNA VE against SARS-CoV-2 infection was 88% in the IC group (N=17,043) versus 94% in the overall study population [20]. In all of these real-world studies conducted to date, the IC sample sizes were relatively small (<2,000 patients), except that of Chodick et al. [17] and Young-Xu et al [20]; also these studies analyzed IC conditions in aggregate or separately from the IC group (e.g., contained a cancer patient group or a chronic kidney disease group). Additionally, definitions of IC and VE differed, as well as study periods, countries, patient demographics, follow-up durations, and data sources. To briefly summarize, mRNA VE against SARS-CoV-2 infection, symptomatic illness, and COVID-19-related hospitalization for IC patients in these existing studies has been found to be consistently lower than observed in the general populations, ranging from 63% to 90% [13-20]. Although our study was unable to determine the VE of BNT162b2 among IC patients, the incidence rates of COVID-19 we observed can be used to generate a directional estimate of VE using the efficacy seen in clinical trials as a benchmark. For example, among the individuals who were IC (N=212,945), if the VE had been 95% as was seen among participants in the BNT162b2 RCT [5], we would have identified ∼125 (=374 × 0.06% / 0.18%) breakthrough infections. Instead, we observed 374, which is approximately 3 times greater than expected. This would correspond to an extrapolated VE of 85% ((=1 -0.18% / 0.06% x (1-95%)). This simplified calculation does not account for differences between IC and non-IC cohorts that may be associated with the risk of breakthrough infections (eg, older age, prevalence of comorbidities, longer risk interval post-vaccination), hence should be interpreted only for purposes of discussing our findings as directionally consistent with some of the existing studies with IC populations [18-20].

The findings for the non-IC cohort in this study are relatively consistent with the VE found for the BNT162b2 RCT population [5], as they relate to the proportion of breakthrough infections over the median follow-up time (2 months in the RCT; 99 days/∼3.3 months in this study). They additionally highlight the greater relative risk of those who are IC for breakthrough infections. As of August 30, 2021, the CDC reported that approximately 173 million people had been fully vaccinated against COVID-19 in the US; also by this date, a total of 12,908 patients had been reported hospitalized (N=10,471) or died (N=2,437) from a COVID-19 vaccine breakthrough infection to the CDC [33]. This is equivalent to 0.0075% of the 173 million people fully vaccinated. These data are generally consistent with our findings among the overall study population (N=1,277,747), in which 124 were hospitalized (with 2 inpatient deaths) for breakthrough infections, which is equivalent to 0.0097%. Such real-world data underscore the effectiveness of vaccines to prevent symptomatic/severe COVID-19 illness regardless of IC status.

Among the IC study cohort, those who were older (age ≥65 years) had an incidence rate of COVID-19 vaccine breakthrough infections of 1.50 per 100 person-years, which was approximately double that of those <65 years of age (0.71 per 100 person-years). Although many of the IC groups for those aged ≥65 years had small sample sizes, those with solid malignancy, rheumatologic/inflammatory conditions, other immune conditions, kidney disease, hematologic malignancies, and >1 IC condition had higher incidence rates of breakthrough infections than among those <65 years of age with the same corresponding IC conditions. The incidence rate of breakthrough infections in the non-IC older subgroup was also approximately double that of the non-IC population <65 years of age (0.65 vs. 0.31 per 100 person-years). The real-world studies of Yelin et al., Whitaker et al., Chodick et al., and Dagan et al. have also reported modest decreases in COVID-19 vaccine protection among older patient groups [15-19]. Our study findings further emphasize the need for augmented prevention and protective measures, particularly for older adults with IC conditions, as well as provide novel insights to help delineate the risk associated with specific IC conditions.

The findings of this study should be interpreted in the context of its limitations. First, COVID-19 vaccination status captured in administrative claims-based data sources may not be comprehensive and some BNT162b2 vaccinated individuals may have been missed, especially with the code-specific definitions of vaccination status utilized in this study. Furthermore, an unvaccinated study group was not included in the analyses, because people who are unvaccinated could not be reliably identified in the administrative claims-based data source. We utilized the ICD-10 code U071 on a healthcare claim to identify a breakthrough infection, and thus asymptomatic/minimally symptomatic breakthrough infection cases that may have been only identified with a RT-PCR test may have been missed, in addition to those individuals who did not seek care for COVID-19 illness. Although we included multiple types of IC conditions in this study, the sample sizes of some IC condition groups were small, especially bone marrow transplant recipients. Additionally, the broad algorithm used to identify individuals with IC conditions may have led to some individuals being categorized as IC when they actually were not. Laboratory test results, if they were available in the data source, may have helped in identifying more precisely individuals who were IC and further study is warranted using other data sources that contain such information. The majority of the study period encompassed the time prior to the dominance of the delta variant in the US, therefore the incidence rates of breakthrough infections reported herein may not generalize beyond the study period. This descriptive study did not adjust for potential differences between the IC and non-IC cohorts, such as the prevalence of comorbidities known to increase the risk for severe COVID-19 (e.g., type 2 diabetes, obesity) and frequency of healthcare utilization.

## Conclusions

To our knowledge, this study contained the largest population of fully vaccinated IC individuals to date in whom COVID-19 vaccine breakthrough infections have been evaluated. The study findings show that breakthrough infections are rare but are more common and severe in people with certain IC conditions. They support FDA authorization and CDC recommendations to offer a 3^rd^ vaccine dose to increase protection among IC individuals [22,23], and the need for vigilant efforts to maximize vaccine uptake among the IC, especially in the context of waning duration of protection and emerging SARS-CoV-2 variants. Moreover, our findings suggest that breakthrough infections can occur regardless of active treatment status in the IC and that there may be additional vulnerable IC groups that could benefit from increased protection. This study advances the understanding of post-vaccination outcomes across multiple IC condition groups in a real-world setting and may help healthcare providers in the decision-making process when vaccinating and treating patients at high-risk for COVID-19.

## Supporting information

STROBE Checklist

Sterling IRB, Boston, Maine waived ethical approval for this work

## Data Availability

Data generated or analyzed during this study are available upon request.

## Funding statement

This study was sponsored by Pfizer Inc.

## Disclosure statement

M Di Fusco, MM Moran, A Cane, D Curcio, F Khan, D Malhotra, A Miles, D Swerdlow, J McLaughlin, and JL Nguyen are employees of Pfizer. A Surinach is an employee of Genesis Research, which has received consulting fees from Pfizer.

## Authorship contributions

All named authors meet the International Committee of Medical Journal Editors (ICMJE) criteria for authorship for this article. All authors contributed to study conception and design, data acquisition, analysis, and interpretation, drafting and revising of the manuscript, and have given their approval for this manuscript version to be published.

## Acknowledgements

The authors acknowledge Luis Jodar and Florin Draica (Pfizer employees) and Mallika Viswanath and Rebecca Nock (HealthVerity employees) for specific contributions to this research project. Editorial support was provided by Jay Lin and Melissa Lingohr-Smith at Novosys Health and was funded by Pfizer.

## Data availability statement

Data generated or analyzed during this study are available upon request.

## Notes

### Author Declarations

This study was deemed exempt from Institutional Review Board (IRB) review pursuant to the terms of the US Department of Health and Human Service's Policy for Protection of Human Research Subjects at 45 C.F.R. 46.104(d); category 4 exemption (Sterling IRB, Boston, Maine waived ethical approval for this work).

