## Supplementary material for "Evaluation of COVID-19 vaccine breakthrough infections among immunocompromised patients fully vaccinated with BNT162b2": Online Supplementary Material

**Supplementary Table 1. ICD-10 codes indicating immunocompromising diseases to identify immunocompromised cases**

| **IC Condition** | **ICD-10 Code** |
| --- | --- |
| HIV/AIDS ^a^ | B20-B24 |
| Solid malignancy |  |
| Organ/system malignant tumors | C00-C07; C11-C19; C22-C80; Z85 |
| Neuroendocrine tumors | C7A; C7B; D3A |
| Neoplasms of uncertain behavior | D00-D49 |
| Bone marrow transplant | Z94.81 |
| Organ transplant |  |
| Complications of transplanted organ | T86 |
| Organ transplant status | Z94 except Z94.81; Z98.85 |
| Rheumatologic or other inflammatory condition |  |
| Sarcoidosis | D86 |
| Amyloidosis NOS | E85 |
| Familial Mediterranean fever | E85.0; M04 |
| Amyloidosis NEC | E85.1; E85.3; E85.8 |
| Multiple sclerosis | G35 |
| Other CNS demyelination | G36; G37.1; G37.3; G37.8; G37.9 |
| Acute infective polyneuritis | G61.0; G61.9 |
| Acute myocarditis | I40 |
| Polyarteritis nodosa and other | M30 |
| Allergic alveolitis/pneumonitis NOS | T78.40; J67.9 |
| Other alveolar pneumonopathy | J84.01; J84.02; J84.09 |
| Enteritis and colitis | K50-K52 |
| Lupus erythematosus | L93.0; L93.2; M32 |
| Diffuse connective tissue disease | L94; M35.8; M35.9 |
| Arthropathy with infection | M12.9; M01.X0; M02.10 |
| Crystal arthropathies | M11 |
| Rheumatoid arthritis/inflammatory polyarthropathy | M05-M14 |
| Inflammatory spondylopathies | M46 |
| Polymyalgia rheumatica | M31.5; M35.3 |
| Chronic inflammatory demyelinating polyneuropathy | G61.81 |
| Immune thrombocytopenic purpura | D69.3 |
| Primary immunodeficiency |  |
| X-linked agammaglobulinemia | D80.8 |
| Common variable immunodeficiency | D83.1; D83.2; D83.8; D83.9 |
| IgA deficiency | D80.2 |
| IgG sub-class deficiency | D80.3 |
| Severe combined immunodeficiency | D81.1 |
| Di George syndrome | D82.1 |
| Wiskott-Aldrich | D82.0 |
| Ataxia telangiectasia | G11.3 |
| Interferon-gamma/Interleukin 12 axis deficiencies | D84.89 |
| Persistent complement, properdin or Factor B deficiency | D84.1 |
| Received eculizumab for ≥14 days during the baseline period |  |
| Chronic granulomatous disease | D71 |
| Chediak-Higashi | E70.330 |
| Leukocyte adhesion deficiency | D72: Genetic anomalies of leukocytes |
| Myeloperoxidase deficiency | D72.89: Other specific disorders of WBCs |
| Other immune conditions |  |
| Disorders of immune mechanism | D89 |
| Neutropenia | D70 |
| Functional disorders of neutrophils | D71 |
| Genetic anomalies of leukocytes | D72.0 |
| Decreased leukocyte count | D72.81 |
| Leukocyte disease NEC | D72.89 |
| Leukocyte disease NOS | D72.9 |
| Myelofibrosis | D75.81 |
| Blood diseases NEC | D47.4; D75.89; D75.9; D89.2 |
| Blood diseases NOS | D75.9; D75.89 |
| Immunologic findings NEC | R76; R83.4-R87.4; R89.4 |
| Nonspecific immune findings NEC and NOS | R76; R83.4-R87.4; R89.4 |
| Sickle cell disease | D57 |
| Asplenia | Q89.01 |
| Psoriatic arthritis | L40.52 |
| Kidney condition |  |
| Chronic kidney disease | A18.11; A52.75; B52.0; C64.x; C68.9; D30.0x; D41.0x-D41.2x; D59.3; E08.2x; E09.2x; E10.2x; E10.65; E11.2x; E11.65; E13.2x; E74.8; I12.xx; I13.0; I13.1x; I13.2; K76.7; M10.3x; M32.14; M32.15; N01.x-N08.x; N13.1; N13.1x-N13.39; N14.x; N15.0; N15.8; N15.9; N16; N17.x; N18.1-N18.5; N18.8; N18.9; N19; N25.xx; N26.1; N26.9; O10.4xx; O12.xx; O26.83x; O90.89; Q61.02; Q61.1x-Q61.8; Q26.0-Q26.39; R94.4 |
| End stage renal disease | N18.6 AND on dialysis (any type): Z99.2; Z49; Z9115; Z4931; OR Z4901 |
| On hemodialysis | Any individual with the ESRD codes above and ≥1 hemodialysis procedure session during the baseline period identified by at ≥1 of the following codes: Z49.31; Z49.32; I953; A4680; A4690; A4706-A4709; A4730; A4740; A4750; A4755; A4802; A4870; A4890; A4918; E1520; E1530; E1540; E1550; E1560; E1575; E1580; E1590; E1600; E1610; E1615; E1620; E1625; E1636; G0365; G0392; G0393; G8081; G8082; G8085; S9335; 90935; 90937; 90940; 93990; 36800; 36810; 36815 |
| On peritoneal dialysis | Any individual with the ESRD codes above and ≥1 peritoneal dialysis procedure session during the baseline period identified by ≥1 of the following codes: Z49.02; 90945; 90947 |
| Hematologic malignancy |  |
| Lymphatic and hematopoietic tissue malignancy | C81-C83; C88-C96 |

AIDS: Acquired immunodeficiency syndrome; CNS: Central nervous system; HIV: human immunodeficiency virus; ICD-10: International Classification of Diseases, 10th Revision; NEC: Necrotizing enterocolitis; NOS: Not otherwise specified; WBCs: White blood cells.
^a^ Excluded asymptomatic HIV code of ICD-10: Z21.

**Supplementary Table 2. Immunosuppressive medications to identify immunocompromised cases**

| **Immunosuppressive Medication Class** | **Medications** |
| --- | --- |
| Chemotherapeutic agents | Aldesleukin; Alemtuzumab; Altretamine; Amifostine; Arsenic trioxide; Asparaginase; Azacitidine; Bendamustine hydrochloride; Bevacizumab; Bexarotene; Bortezomib; Brentuximab vedotin; Busulfan; Cabazitaxel; Capecitabine; Carboplatin; Carfilzomib; Carmustine; Cetuximab; Chlorambucil; Cisplatin; Cladribine; Clofarabine; Cyclophosphamide; Dacarbazine; Dactinomycin; Dasatinib; Daunorubicin citrate liposome; Decitabine; Denileukin diftitox; Docetaxel; Etoposide; Everolimus; Floxuridine; Fluorouracil; Gefitinib; Ifosfamide; Ipilimumab; Ixabepilone; Lomustine; Melphalan; Mercaptopurine; Mesna; Methotrexate; Mitomycin; Mitotane; Nelarabine; Ofatumumab; Oxaliplatin; Paclitaxel; Panitumumab; Pegaspargase; Pemetrexed; Pentostatin; Pertuzumab; Plicamycin; Pralatrexate; Rituximab; Romidepsin; Streptozocin; Temozolomide; Teniposide; Thioguanine; Thiotepa; Trastuzumab; Tretinoin; Vorinostat |
| Immunomodulators | Abatacept; Adalimumab; Alefacept; Anakinra; Auranofin; Aurothioglucose; Azathioprine; Basiliximab; Belatacept; Belimumab; Certolizumab pegol; Cyclosporine; Daclizumab; Denosumab; Eculizumab; Efalizumab; Etanercept; Everolimus; Gold sodium thiomalate; Golimumab; Infliximab; Interferon alfacon-1; Leflunomide; Lenalidomide; Mycophenolate mofetil; Natalizumab; Palifermin; Palivizumab; Pegademase bovine; Pimecrolimus; Sirolimus; Tacrolimus; Thalidomide; Tocilizumab; Ustekinumab |
| Systemic corticosteroids | Dexamethasone; Methylprednisolone; Prednisolone; Prednisone (≥60mg/day); Cortisone; Hydrocortisone |
| Antimetabolites | Mycophenolate mofetil; Mycophenolic acid; Azathioprine |

**Supplementary Table 3. Codes for identifying healthcare prevention seeking behavior anytime during the baseline period**

| **Healthcare Seeking Behavior** | **Current Procedural Terminology-4 Codes** |
| --- | --- |
| Telehealth or telephone visit | G2012; G2010; G2061-G2063; 99205-99205; 99212-99215; 99421-99423; 99441-99443; G0425-G0427; G0406-G0408 |
| COVID-19 laboratory test | 87635; 86318; 86328; 86769; 87426; 86408; 86409; 86413 |
| Influenza vaccine | 90653-90664; 90666-90668; 90724; 90470; 90672; 90673; 90685-90688 |

**Supplementary Table 4. Prevalence of IC conditions and IS medication usage among the IC cohort that received 2 BNT162b2 doses**

|  |  | **% of IC Condition Group** | |
| --- | --- | --- | --- |
| **IC condition group** | **N (% of Total IC Cohort)** | **Usage of IS Medications in Baseline Period** | **With Active Treatment^a^ in Baseline Period** |
| Symptomatic HIV/AIDS (excluding asymptomatic HIV) | 2184 (1.0) | 58 (2.7) | 5 (0.2) |
| Solid malignancy | 72,282 (32.0) | 4326 (6.0) | 784 (1.1) |
| Bone marrow transplant | 13 (0.01) | 2 (15.4) | 0 (0.0) |
| Organ transplant (excluding bone marrow) | 710 (0.3) | 472 (66.5) | 142 (20.0) |
| Rheumatologic/other inflammatory condition | 37,782 (16.7) | 10,508 (27.8) | 2919 (7.7) |
| Primary immunodeficiency | 3446 (1.5) | 187 (5.4) | 28 (0.8) |
| Other immune condition^b^ | 3959 (1.8) | 408 (10.3) | 106 (2.7) |
| CKD or ESRD | 44,122 (19.5) | 2153 (4.9) | 293 (0.7) |
| Hematological malignancy | 1894 (0.8) | 258 (13.6) | 79 (4.2) |
| IS medication usage ≥14 days during baseline period | 20,659 (9.1) | 20,659 (100) | 2448 (11.9) |
| Antimetabolite usage ≥14 days during baseline period | 696 (0.3) | 696 (100) | 202 (29.0) |
| >1 IC condition | 38,049 (16.9) | 8202 (21.6) | 2468 (6.5) |
| *Overall IC cohort* | 225,796 (100) | 47,929 (21.2) | 9474 (4.2) |

^a^ Active treatment was defined as evidence of medication usage ≤14 days before a 1^st^ BNT162b2 dose, except for hematological malignancy, where active treatment status was defined as evidence of any IS medication (including radiotherapy) usage within 6 months before a 1^st^ BNT162b2 dose.

^b^ Other immune conditions are described in Supplementary Table 1.

AIDS: Acquired immunodeficiency syndrome; CKD: Chronic kidney disease; ESRD: End stage renal disease; HIV: human immunodeficiency virus; IC: Immunocompromised; IS: Immunosuppressive

**Supplementary Table 5. Characteristics of organ transplant recipients that received 2 BNT162b2 doses**

|  | **N=710** |
| --- | --- |
| *Age (years), mean (SD)* | 53.3 (16.1) |
| Median (Q1-Q3) | 56 (43-63) |
| <65 years, N (%) | 569 (80.1) |
| ≥65 years, N (%) | 141 (19.9) |
| *Sex, N (%)* |  |
| Female | 305 (43.0) |
| Male | 395 (55.6) |
| Unknown | 10 (1.4) |
| *Month of receipt of 1^st^ vaccine dose (cumulative), N (%)* |  |
| December or January | 95 (13.4) |
| February | 221 (33.1) |
| March | 565 (79.6) |
| April | 667 (93.9) |
| May | 698 (98.3) |
| June | 710 (100) |
| July | 710 (100) |
| *Transplant ≤100 days prior to receipt of 1^st^ BNT162b2 dose, N (%)* | 507 (71.4) |
| *Transplant >100 days prior to receipt of 1^st^ BNT162b2 dose, N (%)* | 203 (28.6) |
| *Follow-up duration (days), mean (SD)* | 106.1 (38.0) |
| Median (Q1-Q3) | 110.5 (83-128) |

SD: Standard deviation
