## Supplementary material for "Evaluation of COVID-19 vaccine breakthrough infections among immunocompromised patients fully vaccinated with BNT162b2": Sterling IRB, Boston, Maine waived ethical approval for this work

SterlingIRB
SAFEGUARDING YOUR RESEARCH

**TYPE OF REVIEW – EXEMPTION FROM IRB REVIEW DETERMINATION**

| | |
|---|---|
| Determination Date: | October 12, 2021 |
| IRB ID: | 9339-MDiFusco |
| Protocol: | Coronavirus Disease 2019 (COVID-19) Vaccination and Breakthrough Infections Among Persons with Immunocompromising Conditions in the United States |
| Sponsor: | Pfizer Inc. |
| Principal Investigator: | Manuela Di Fusco, MS |

Sterling IRB is in receipt of submission materials for the above-referenced study.

**Items Reviewed:**

- Exemption or Non-Human Subjects Research Determination Request
- Protocol C4591035 COVID19 NIS IC v1.0 FINAL

Based on the information available to the IRB, the Sterling IRB Chairman (or designee) has determined that:

The above-listed study is exempt from IRB review pursuant to the terms of the U.S. Department of Health and Human Service's Policy for Protection of Human Research Subjects at 45 C.F.R. §46.104(d).

Sterling IRB has determined that the following exemption category(ies) applies:

- Category 4 Exemption (DHHS)

Sterling IRB's exemption determination is based on the study-related information available to Sterling IRB as of the determination date listed above. Should any changes be made to the study subsequent to Sterling IRB's determination, this determination is no longer applicable.

*As the project applicant you are responsible for following all policies of Sterling IRB as described in the Exemption or Non-Human Subjects Research Determination Request Submission Agreement which you accepted with project submission. It is your responsibility to ensure this project is conducted in accordance with applicable regulations (local, state and federal) as well as any requirements established by the IRB at the time of the review determination. Refer to the Investigator Handbook at www.sterlingirb.com for details of these responsibilities.*

The Board will be apprised of this determination.
